# Developmental trajectories of suicidality and lifestyle factors across adolescent severity profiles

**DOI:** 10.64898/2026.08.19.26360854

**Authors:** Maddison Crethar, Daniel F. Hermens, Taliah Prince, Lia Mills, Nicholas Brander-Peetz, Amanda Boyes

## Abstract

**Background:** Adolescent suicide is a leading cause of death in Australia, arising from multiple determinants. Psychological distress, lifestyle behaviours and socioeconomic factors are associated with adolescent suicidality. Existing research has predominantly employed cross-sectional designs, limiting our understanding of how these factors interact over time. Longitudinal and data-driven approaches are needed to help identify the factors associated with the emergence of suicidality throughout adolescence.

**Method:** Participants aged 12–17 years completed longitudinal measures of suicidal ideation, psychological distress, sleep quality, mindfulness, physical activity, eating habits, and social connectedness. Subgroups were determined via hierarchical cluster analysis, based on average scores across later timepoints (9–15). ANOVA and pairwise effect size calculations were used to compare clusters across variables, and their preceding developmental trajectories were examined using generalised additive mixed models (across earlier timepoints; 1–8). Clusters were also compared on self-reported wellbeing, long-term suicidality, and socioeconomic status.

**Result:** Three clusters characterised by low-, moderate-, and high-severity of suicidal ideation and psychological distress, and poorer sleep, social connectedness, physical activity, mindfulness, and eating habits were identified. Across earlier timepoints, the high-severity group showed consistently elevated suicidality and deteriorating wellbeing and lifestyle scores.

**Conclusion:** Youth with high levels of suicidality had greater psychological distress, lower wellbeing, lower socioeconomic status, and poorer lifestyle behaviours. This subgroup was also found to have poorer scores on wellbeing and lifestyle factors in their early adolescence. Findings highlight the importance of early, preventative interventions targeting both mental health and lifestyle factors to reduce suicidality in adolescents.

## INTRODUCTION

Suicide is the leading cause of death of Australian adolescents (AIHW, 2025). For every life lost to suicide, approximately 20 more individuals have attempted suicide (AIHW, 2025). Suicidality tends to increase in adolescence, typically from around age 15 onwards (AIHW, 2025; Glenn et al., 2020). Risk factors for suicidal thoughts and behaviours (STBs) are multidimensional, including neurobiological, epigenetic, genetic, environmental and psychosocial factors (Glenn et al., 2020; Goldstein & Franzen, 2022). Experiences of stressors and impulsivity contribute to fluctuations of suicidality during adolescence (Platt, Kadosh & Lau, 2013). Approximately 17.9% of adolescents aged 11-18yrs experience psychological distress (Shawon et al., 2023). Greater psychological distress (Shawon et al., 2023), lower socioeconomic status (SES), older age, family dysfunction, and violence have been linked with suicidality (Madigan & Daly, 2023). Additionally, these risk factors impact wellbeing. For instance, low SES may reflect reduced living standards and access to resources, which are associated with lower wellbeing (Cosma et al., 2020). Psychological distress, SES, and wellbeing represent interconnected pathways through which multiple factors converge to influence adolescent suicidality. Most adolescents with suicidal ideation do not engage in suicidal behaviours; however, among those who do, the transition often occurs within one year (Nock et al., 2013). There is a need to understand varying levels of severity to inform interventions for at-risk adolescents; however, many studies rely on cross-sectional data.

Lifestyle factors are potential targets for interventions among at-risk adolescents (Xiao, Romanelli & Lindsey, 2019). Data from the Australian national Youth Risk Behaviour Survey (2017) of 14,506 adolescents aged 12 to 18 years revealed that consistently engaging in health-promoting behaviours - such as healthy eating, physical activity, and sleep - contributes to a reduced risk of suicidality (Xiao et al., 2019). Adolescents experience rapid changes in their sleep-wake cycles and circadian regulation (Goldstein et al., 2022). Environmental factors such as increased screen time compound these biological shifts, contributing to poor sleep quality (LeBourgeois et al., 2017). Additionally, prior studies have linked poor sleep quality to increased risk of suicidality (Goldstein et al., 2022; Gowin et al., 2024; Pfledderer, Burns & Brusseau, 2019). The relationship between sleep and suicidality in adolescents appears to have a U-shape association, where too little or too much sleep is positively related to suicidality (Xiao et al., 2019).

Mindfulness refers to present-focused, awareness, and is associated with lower rumination and improved emotional regulation (Faura-Garcia, Orue & Calvete, 2021; Gu, Fang & Yang, 2023). During developmental periods characterised by heightened impulsivity and emotional reactivity, mindfulness practices may promote regulatory capacities and adaptive coping (Amada & Shane, 2018). A study of 609 Korean adolescents found a negative correlation between mindfulness and suicidal ideation (Lee, 2023). However, most research in this area has been cross-sectional, based on interventional trials with small samples, and has shown mixed results depending on how suicidal ideation is assessed (Forkmann, Brakemeier, Teismann, Schramm & Michalak, 2016). Recent findings also indicate adolescents with diminished levels of physical activity demonstrate elevated suicidality and psychological distress (Huo, Yang, Yang & Chen, 2024; Lee & Lee, 2024). However, the association between suicidality and exercise appears non-linear and bidirectional (Grasdalsmoen et al., 2020), and given the historical reliance on adult populations, limited sample sizes, and cross-sectional methods, further research is needed to establish generalisability.

Several large-scale studies link unhealthy eating patterns and suicide-related outcomes in adolescents (Lee et al., 2022; Liu, Jia, Li, 2022; Khan, Ahmed & Burton, 2020). Low vegetable intake and frequent consumption of fast food or caffeinated beverages were linked to increased suicide risk, though methodological limitations such as non-standardised temporal measurement suggest further research is needed to confirm the reliability of these findings across time. Social connectedness is a well-established protective factor against adolescent suicidality (Miller, Esposito-Smythers & Leichtweis, 2015; Xiao & Lindsey, 2021). Strong peer and school relationships buffer the effects of poor mental health on suicidal ideation (Miller et al., 2015; Simcock et al., 2021). However, the protective effect of connectedness may be reduced among adolescents with prior suicidal behaviour or a history of victimisation (Arango et al., 2023), suggesting that connectedness may be more effective as a preventive rather than remedial factor.

While research has examined associations between suicidality and individual lifestyle factors, the predominance of approaches based on single variables, cross-sectional analyses, or quasi-longitudinal methods has limited our understanding of how these factors interact over time. This represents a significant gap, as adolescent suicidality emerges from the dynamic interplay of multiple risk and protective factors rather than from isolated variables (Glenn et al., 2020; Pfledderer et al., 2019). The objective of this study was therefore to analyse suicidality, psychological distress, and lifestyle factors from a sample of mid-adolescents undertaking a longitudinal study, to determine various subgroups, and to then retrospectively examine whether these groups have different longitudinal trajectories from early adolescence. By clustering adolescents across multiple dimensions simultaneously, this approach aims to identify distinct severity profiles and inform targeted prevention and intervention strategies.

## METHODS

### Participants and Design

Data were acquired between 2018 and 2025 as part of the Longitudinal Adolescent Brain Study (LABS), which tracks neurobiological and psychological changes in Australian adolescents from the age of 12 to 17 years at four-month intervals, with up to 15 timepoints. Participants were recruited through local media, community services, and schools. Informed consent was obtained from the parent/guardian and young person prior to participation. Exclusion criteria included major neurological disorder, intellectual disability, medical illness (non-psychiatric), or sustained a head injury with loss of consciousness for over 30 minutes. As at 28 January 2025, 179 participants were enrolled in LABS. In the present sample (N=98), participants were aged between 15.3 and 17.9 years (*M* = 16.2; 56% female; 21.43% had a mental health diagnosis), with clusters derived from timepoints 9-15 to assess profiles in mid-adolescence. Attendance was varied (refer to Figure 1 for a breakdown by timepoint). If a participant missed a timepoint, they were able to rejoin the study at the next fixed timepoint, at four-monthly intervals (from their first timepoint).

**Figure 1:**
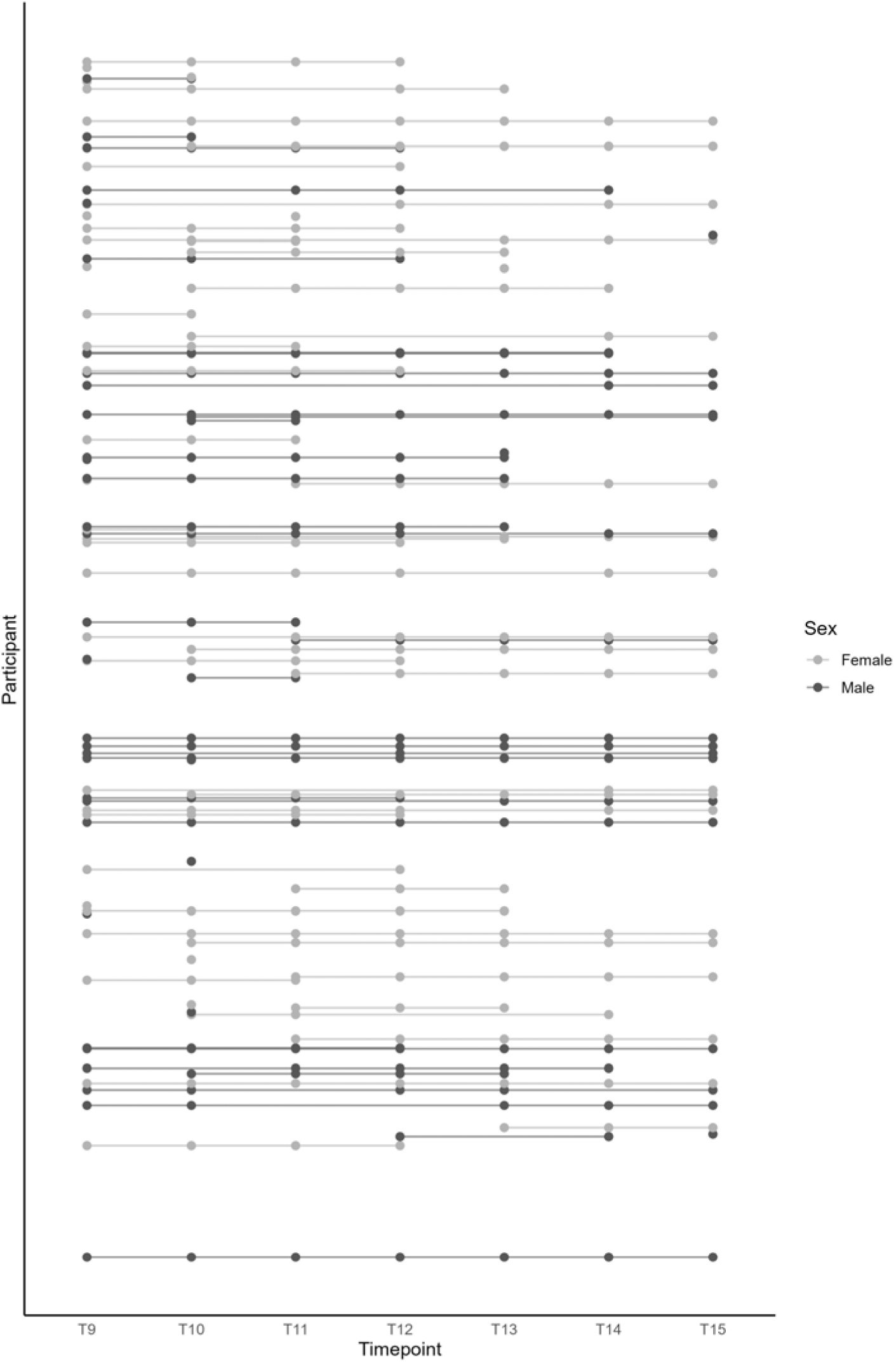
Total Assessment Count: Attended Timepoints (T9-15) by Participants (N=98)

### Measures

#### Suicidality

The Suicidal Ideation Attributes Scale (SIDAS) is a five-item scale measuring suicidal ideation severity in the past month (van Spijker et al., 2014). Responses are rated on an 11-point Likert scale (0 = *never,* 10 = *always*). Scores range from 0-50, with higher scores indicating greater severity (Torok et al., 2022). The SIDAS has high validity and reliability (van Spijker et al., 2014) and has been validated for use with 16–25-year-olds (lorfino et al., 2017). The Youth Risk Behaviour Survey (YRBS) includes four items assessing the presence of STBs within the past year. Items 1 and 2 assess ideation and planning on a dichotomous scale (0 = *no*, 1 = *yes*); item 3 assesses number of attempts on a scale from 0 (*0 times*) to 4 (*6 or more times*); item 4 assesses if an attempt required medical treatment on a dichotomous scale (0 = *I did not attempt* or *no*, 1 = *yes*). Item scores were summed to create a total score ranging from 0 to 7. Higher scores indicate greater suicidality severity in the past year. The YRBS items have demonstrated good convergent and discriminant validity in adolescent samples (May & Klonsky, 2011). Previous research in this cohort demonstrated YRBS total score’s utility in assessing adolescent mental health, capturing history of suicidal behaviours (Beaudequin et al., 2020).

#### Psychological distress

The K10 is a 10-item self-report questionnaire measuring non-specific symptoms of psychological distress, where respondents report how often they have experienced such symptoms in the previous 30 days (Kessler et al., 2002). Responses are rated on a 5-point Likert scale (1 = ***n****one of the time*, 5 = ***a****ll of the time*). Scores range from 10 (low levels of distress) to 50 (severe levels). The K10 is widely used in Australian and international health instruments and has been validated among adolescent samples (Jakobsen, Larsen & Horwood, 2017).

### Eating Habits

The Food Frequency Questionnaire (FFQ) is derived from the HBSC survey and includes six items (Roberts et al., 2009). It captures the frequency of consuming fruit, vegetables, sweets, and sugar-sweetened soft drinks, along with breakfast on weekdays and weekends. Four items ask: “How many times a week do you usually eat or drink…?” with response options scored on a 7-point Likert scale (0 = *never*, 6 = *every day, more than once*). Two items assess breakfast consumption; one for weekdays (scored 0-5) and one for weekends (scored 0-2). Items reflecting unhealthy behaviours are reverse scored. Total scores range from 0-31, with higher scores indicating healthier eating habits. The FFQ has been validated for use with 11-12-year-olds (Vereecken, Rossi, Giacchi & Maes, 2008).

### Mindfulness

The Mindful Attention Awareness Scale for Adolescents (MAAS-A) includes 14 items that capture levels of mindfulness (Brown, West, Loverich & Biegel, 2011). Items are scored on a 6-point Likert scale (1□=□a*lmost always,* 6□=□a*lmost never),* with higher scores indicating higher trait mindfulness (de Bruin, Zijlstra, van de Weijer-Bergsma & Bögels, 2011). The MAAS-A is an adapted measure of the adult version (MAAS) and has been validated for use in populations aged 11-17 years (Brown et al., 2011; de Bruin et al., 2011).

### Sleep

The Pittsburgh Sleep Quality Index (PSQI) consists of 19 items and seven subscales of sleep over the past month, including: quality, latency, duration, habitual efficiency, disturbance, medication usage, and daytime dysfunction (Buysse, Reynolds, Monk, Berman & Kupfer, 1989). Responses are scored on a 4-point Likert scale ranging from 0-21, with scores above five being associated with poor sleep quality. Two items were adjusted to enhance applicability to adolescent populations. Item seven, assessing trouble with “staying awake while driving, eating meals, or engaging in social activity”, was altered by replacing “driving” with “studying”. Item ten was removed as it relates to bed partners or roommates. The PSQI has been validated in adolescents (de la Vega et al., 2015).

### Social Connectedness

The social connectedness scale (SCS) is a 15-item questionnaire measuring self-reported sense of closeness and belonging to others across familial, school, peer, and community relationships (Lee & Robbins, 1995). Participants rate their response on a 6-point Likert scale (1 = s*trongly disagree*, 6 = s*trongly agree*). Example item: “I feel disconnected from the world around me”. Scores range from 15-90, with higher scores indicating higher social connectedness. The SCS has been validated with adolescents (Grieve, Indian, Witteveen, Tolan & Marrington, 2013).

### Physical Activity

This instrument was derived from the WHO health behaviour in schoolchildren (HBSC) survey (Roberts et al., 2009). It consists of three items relating to moderate-to-vigorous physical activity (MVPA) and vigorous physical activity (VPA). MVPA item (1): “Over the past 7 days, on how many days were you physically active for a total of at least 60 minutes per day?” (range: 0-7). VPA item (1): “How often do you usually exercise in your free time so much that you get out of breath or sweat?” (responses range from 0 = *never* to 6 = e*very day*). VPA item (2): “How many hours a week do you usually exercise in your free time so much that you get out of breath or sweat?” (responses range from 0 = *none* to 6 = *about 7 hours or more)*. Scores are summed to produce a total score ranging from 0 to 19, with higher scores indicating greater levels of physical activity. The MVPA and VPA items have been respectively validated in samples of 15–17-year-olds (Ridgers, Timperio, Crawford & Salmon, 2012) and 13–15-year-olds (Booth, Okely, Chey & Bauman, 2001).

### Wellbeing

The COMPAS-W assesses self-reported wellbeing across 26 items (Gatt et al., 2014). It comprises of six constructs (Composure, Own-worth, Mastery, Positivity, Achievement, and Satisfaction) that contribute to an overall measurement of wellbeing (Gatt et al., 2014). Response are scored on a 5-point Likert scale (1 = *strongly disagree,* 5 = *strongly agree*). Total scores range from 26-130 and higher scores represent greater wellbeing. The COMPAS-W has been validated for use with adolescents (Lam et al., 2024).

### Socioeconomic Status

Participant postcodes were recoded into socioeconomic (SES) quintiles using the Index of Relative Socioeconomic Advantage and Disadvantage (Australian Bureau of Statistics, 2021).

### Data Analyses

Statistical analyses were performed using IBM SPSS Statistics version 29 and R version 4.4.1. Cluster analyses utilised data from timepoints 9–15 (*M* = 16.20; *SD* = 0.7) to group individuals into suicidality severity profiles. Aggregated means were created for each participant for each variable, across timepoints 9–15. This approach accommodated variability in the number of timepoints completed across participants and prevented case-level missing data entering the cluster analysis. Mean scores were standardised, with scores inverted for SIDAS, K10, and PSQI to ensure directional alignment across variables (before inversion, higher scores on the SIDAS, K10, and PSQI represented ‘worse’ scores, while higher scores on the FFQ, PA, MAAS-A and SCS represented ‘better’ scores). Outliers beyond Z-score values of -3.0 or +3.0 were winsorised to ±3.0 to reduce the impact of extreme scores (Hermens, Redoblado Hodge, Naismith, Kaur, Scott & Hickie, 2011). The number of outliers did not exceed 10% for any variable.

A hierarchical cluster analysis using Ward’s method of minimum variance with a squared Euclidean distance measure was conducted to explore patterns of suicidality, psychological distress, and engagement in lifestyle factors among subgroups of individuals. This cluster analysis technique has been used in studies of lifestyle factors (López-Gil, Brazo-Sayavera, García-Hermoso, Camargo & Yuste Lucas, 2020) and suicide (Sinyor, Tan, Schaffer, Gallagher & Shulman, 2016), with hierarchical clustering recommended for smaller datasets. In line with previous research (Xiao et al., 2019), variables included were the SIDAS, K10, PSQI, FFQ, MVPA, MAAS-A and SCS. Visual inspection techniques (agglomeration schedule, dendrogram, and scree plot) were used to examine cluster characteristics.

One-way between-subjects analyses of variance (ANOVA) examined differences across clusters on all variables, with Cohen’s d calculated to quantify pairwise effect sizes. A discriminant function analysis (DFA) determined which variable combinations best distinguished between clusters and predicted group membership. Lastly, group-level trajectories were modelled across clusters using GAMMs in R (‘*mgcv’* package). GAMMs can quantify average developmental trajectories without predefining the expected trajectory shape, while accounting for individual-level variability (Wood, 2017). Separate models were fitted for each outcome using cluster-specific cubic regression splines (k = 4) via REML, following the formula: *outcome ∼ cluster + s(tp, by = cluster, k = 4, bs = "cr") + s(subj_id, bs = "re").* The inclusion of a random intercept for participants accounted for repeated measures within individuals. This approach allowed trajectories to vary non-linearly across clusters while controlling for overall between-cluster differences.

## RESULTS

### Cluster Characteristics

The agglomeration coefficients produced by the cluster analysis along with visual inspection of the dendrogram and scree plot revealed a three-cluster solution (*low-severity: n* = 43; *moderate-severity*: *n* = 37; *high-severity*: *n* = 18; see Figures S1-S2). Table S1 displays mean Z-scores (winsorised at ±3.0) and standard deviations for the seven cluster variables, along with the ANOVA-derived F statistics, significance values, and Cohen’s d effect sizes. The cluster characteristics revealed significant between-group differences across the three clusters for eating habits, mindfulness, physical activity, psychological distress, sleep, social connectedness and recent suicidal ideation (<1 month). Table 1 depicts mean scores and standard deviations for demographic and self-report variables across clusters between timepoints 9–15, with ANOVA results displaying between-group differences. Significant between-group differences were detected across the three clusters for sex, socioeconomic status, wellbeing and longer-term STBs (<12 months), with no significant difference detected for age.

**Table 1:** Demographic and Self-Report Characteristics with Standardised Mean (SD) Variable Scores Across Clusters (T9-15)

| Variable | Mean (SD) |  |  | Significance test [p value] | Effect sizes, *d* |  |  |
| --- | --- | --- | --- | --- | --- | --- | --- |
|  | *Low-severity*<br>(*n* = 43) | *Moderate-severity*<br>(*n* = 37) | *High-severity*<br>(*n* = 18) |  | *Low- vs. High-severity* | *Low- vs. Moderate-severity* | *High- vs. Moderate-severity* |
| Sex assigned at birth [f/m] | [18/25] | [24/13] | [12/6] | $\chi^2(2, 361) 13.6 [.001]**$ | - | - | - |
| Age, years | 16.15 (0.76) | 16.19 (0.68) | 16.29 (0.69) | $F(2,358) = 2.7 (.069)$ | -0.19 | -0.06 | 0.15 |
| Socioeconomic Status (SES) | 3.55 (0.88) | 3.63 (0.75) | 3.24 (1.01) | $F(2,358) = 4.5 (.012)*$ | 0.34* | -0.10 | -0.46* |
| Wellbeing (COMPAS-W) | 106.20 (6.20) | 95.45 (7.31) | 83.41 (10.61) | $F(2,358) = 224.0 (<.001)**$ | 2.95† | 1.60† | -1.41† |
| Long term STBs (YRBS) | 0.00 (0.00) | 0.01 (0.06) | 0.58 (0.89) | $F(2,358) = 58.5 (<.001)**$ | -0.65‡ | -0.17 | 1.13† |

*Note.* Cohen's *d* effect sizes (*d*) provided: †Large effect ( $d > 0.8$ ); ‡Medium effect ( $d > 0.5$ ); \*Small effect ( $d > 0.2$ ). ANOVA = analysis of variance. Three instances had missing SR data albeit with available demographic data, resulting in a total *n* difference between sex (361) and other variables (358).

The *low-* and *high-severity* clusters showed the largest differences, with large effect sizes across wellbeing, psychological distress, sleep, mindfulness, social connectedness, physical activity and eating habits (Cohen’s *d* > 0.8). The *high-* and *moderate-severity* clusters also differed significantly across most variables, with eating habits showing the smallest between-group difference (*d* = -0.41). The *low-* and *moderate-severity* clusters differed significantly across most variables, with the exception of recent suicidal ideation (SIDAS; *d* = -0.01) and long-term STB (YRBS; *d* = -0.17), indicating these two clusters were most similar in their suicidality profiles. Table S2 displays the prevalence of YRBS suicide-related behaviours. The *high-severity* cluster had the highest prevalence of STBs, with 50.00% (*n=*9) of participants reporting previously considering suicide; 33.33% (*n=*6) having made a suicide plan and 22.22% (*n=*4) previously attempting suicide. The *low-severity* cluster had the lowest levels of suicidality and psychological distress, and the best sleep, social connectedness, physical activity, mindfulness traits, and eating habits. The *moderate-severity* cluster reported scores between both the *low-* and *high-severity* clusters across all variables except on measures of suicidality, reporting similar scores to that of the *low-severity* cluster. The *moderate-severity* cluster had moderate psychological distress, sleep, social connectedness, mindfulness traits, eating habits, and physical activity. The *high-severity* cluster exhibits the highest levels of suicidality and psychological distress, and the poorest sleep, social connectedness, mindfulness traits, eating habits, and physical activity.

### Discriminant Function Analysis

The seven variables were entered simultaneously as predictors into the DFA. The DFA confirmed profiles by generating two functions to separate the three clusters. The first function accounted for 93.1% of the differences among clusters (Wilks’ λ = .131, χ²(14) = 722.115, *p* < .001). The second explained the remaining 6.9% variance and was also significant (Wilks’ λ = .742, χ²(6) = 105.847, *p* < .001). The structure matrix showed a clear delineation of psychological distress (K10, *r* = .596), sleep (PSQI, *r* = .543), mindfulness traits (MAAS-A, *r* = .536), social connectedness (SCS, *r* = .411), eating habits (FFQ, *r* = .347) and suicidal ideation (SIDAS, *r* = .314) versus function 2 which had high discriminant loadings for suicidal ideation (SIDAS, *r* = .670), mindfulness traits (MAAS-A, *r* = -.623) and eating habits (FFQ, *r* = -.533). Function 1 accounted for the majority of variance and supported the hierarchical clustering technique.

In investigating each predictor’s relative contribution to discriminating groups, the standardised canonical discriminant function coefficients for the first function were examined. Scores indicate the weight of each predictor on maximising group differences. They displayed relatively high correlations for SIDAS (.503) and PSQI (.485). The MAAS-A, SCS, MVPA, FFQ, and K10 had moderate to minimal contributions, with coefficients of .362, .297, .295, .253, and .166. These outcomes demonstrate that SIDAS was the strongest discriminator of group membership, with PSQI showing a similarly large contribution.

Mental health and wellbeing trajectories for timepoints 1-8 were examined. Figure 2 shows the mean SIDAS scores of individuals within each cluster over timepoints 1–8. This demonstrates that the clusters, determined from aggregated means across timepoints 9–15, were consistently low, moderate or high in suicidality scores, respectively, at least from timepoint 2. Of note, these differences were not as apparent at timepoint 1. The error bars indicate greater variability in SIDAS scores for the *high-severity* cluster, compared to the other clusters, across earlier timepoints.

**Figure 2:**
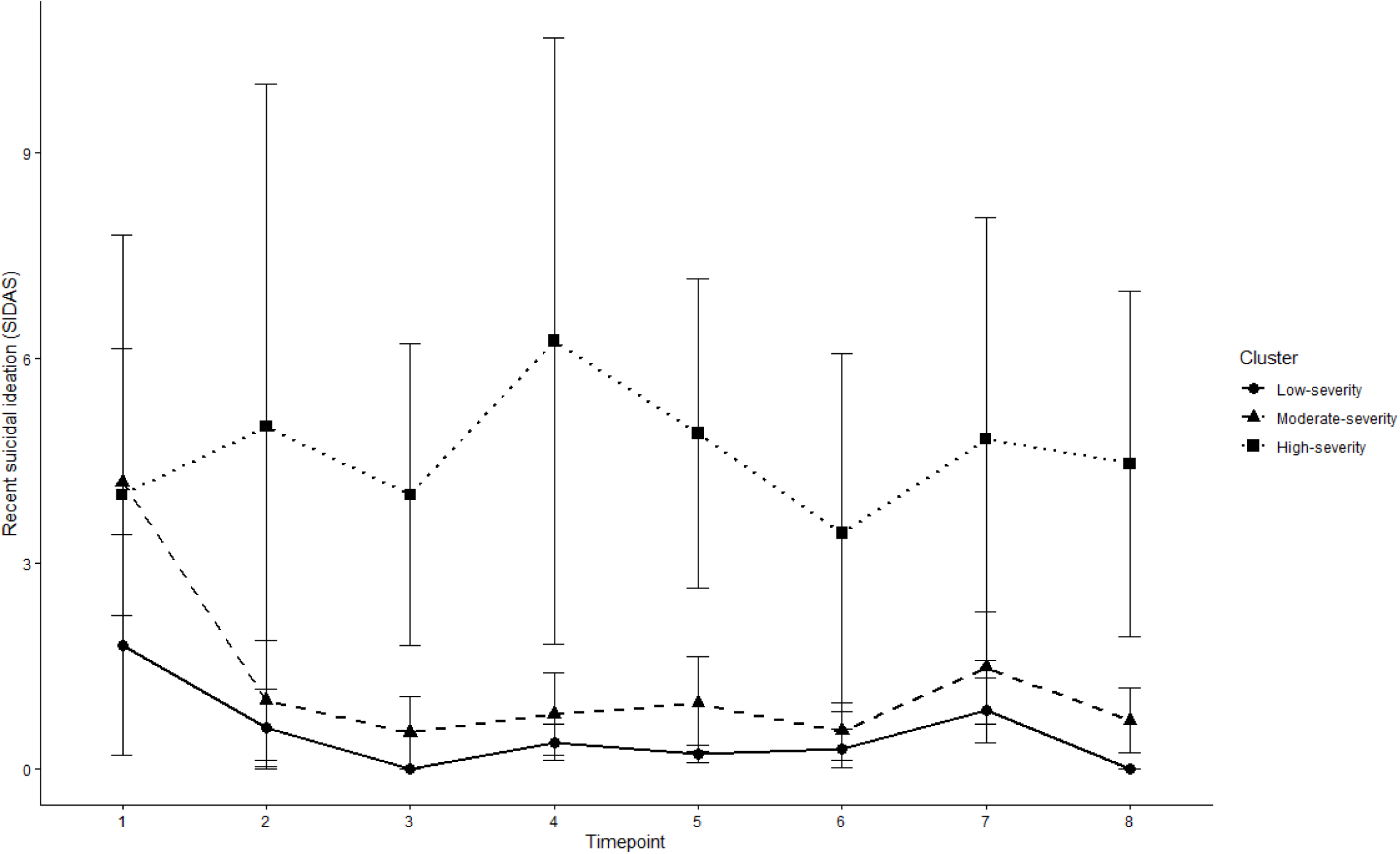
Mean SIDAS scores by cluster group across early adolescence (T1-8) *Note.* Profile of mean SIDAS scores (with standard error bars [1 standard error]) across model variables, arranged by cluster group for timepoints 1 to 8.

To visualise cluster-level trajectories, mean standardised outcome scores were calculated at each timepoint (T1-8) within clusters. Figure 3 displays trajectories for K10, FFQ, MAAS-A, MVPA, PSQI, SCS, and COMPAS-W, across timepoints 1-8. Wellbeing and lifestyle factors varied considerably from timepoint 1 (*M* = 12.6 years) to timepoint 8 (*M* = 15.0 years). The *low-severity* cluster maintained consistently positive standardised scores across all variables. The *moderate-severity* cluster displayed intermediate scores, generally falling between the other two clusters, across all variables and timepoints. The *high-severity* cluster demonstrated the poorest outcomes, with notable deterioration observed around timepoints 3-4, remaining substantially below the other clusters at later timepoints.

**Figure 3:**
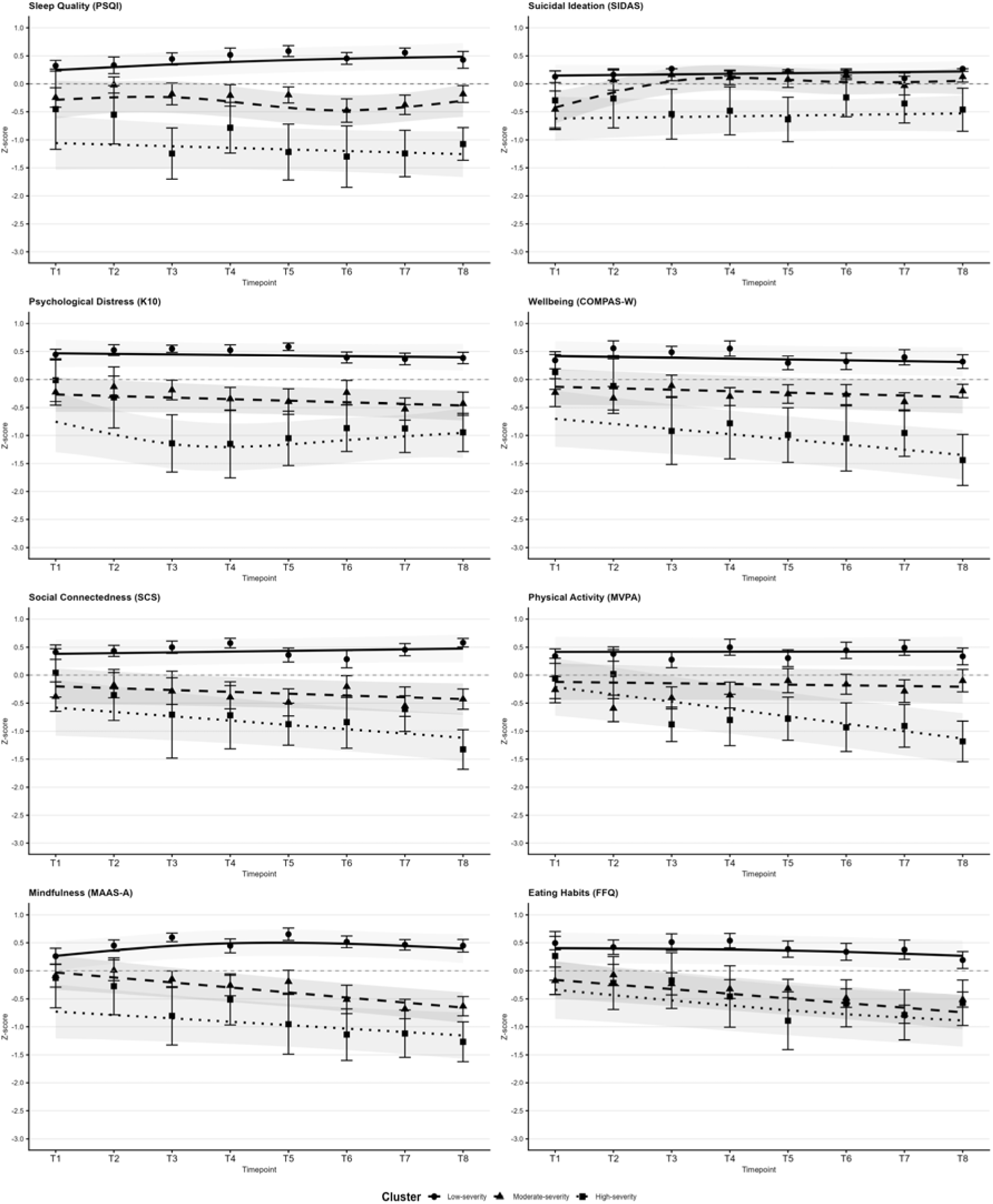
Cluster trajectories across timepoints 1-8 *Note.* Cluster mean z-scores (with standard error bars [±1 SE]) and GAMM-smoothed trajectories across model variables (SIDAS, K10, FFQ, MAAS-A, PA, PSQI and SCS) and COMPAS-W, timepoints 1–8. (a) Recent suicidality (SIDAS), (b) Wellbeing (COMPAS-W), (c) Sleep quality (PSQI), (d) Social connectedness (SCS), (e) Psychological distress (K10), (f) Physical activity (MVPA), (g) Mindfulness (MAAS-A), (h) Eating habits (FFQ). Z-scores were inverted for SIDAS, K10, PSQI to ensure directional alignment across variables (before inversion, higher scores on the SIDAS, K10, and PSQI represented ‘worse’ scores, while higher scores on the FFQ, PA, MAAS-A and SCS represented ‘better’ scores. Outliers beyond Z-score values of -3.0 or +3.0 were winsorised to ±3.0 prior to GAMMs. The number of outliers for the *high-severity cluster* exceeded 10% of cases for SIDAS, SCS, COMPAS-W, K10 and YRBS. GAMM smooths were estimated using cluster-specific cubic regression splines (*k* = 4) fitted via REML

GAMMs confirmed the *high-* and *moderate-severity* clusters had significantly poorer outcomes than the *low-severity* cluster across most variables, consistent with the ANOVA findings (refer to Table S3 for GAMM results). Exceptions were noted for measures of suicidality where the *moderate-severity* cluster did not differ significantly from the *low-severity* cluster (SIDAS, *b* = -0.21, *p* = .098; YRBS, *b* = -0.07, *p* = .563). Trajectory analyses revealed that the *low-severity* cluster remained stable across all timepoints, with no significant smooth terms observed for any outcome. The *high-severity* cluster demonstrated significant change over time for wellbeing (*F* = 8.51, *p* = .004), social connectedness (*F =* 4.76, *p* = .030), physical activity (*F =* 17.00, *p* <.001), eating habits (*F* = 4.25, *p* = .016), and STBs (*F* = 4.40, *p* <.001). Inspection of the model-predicted trajectories indicated these changes reflected progressive decline, with the most pronounced deterioration observed for physical activity and wellbeing. The *moderate-severity* cluster showed significant trajectories for suicidal ideation (*F =* 5.65, *p =* .006), mindfulness (*F =* 18.86, *p* < .001), and eating habits (*F =* 22.82, *p* <.001), with visual inspection indicating deterioration in mindfulness and eating habits over time.

Measures of suicidality showed largely stable trajectories; however, the *moderate-severity* cluster showed a significant non-linear trajectory for recent suicidal ideation (SIDAS; *F =* 5.65, *p =* .006) and the high-severity cluster showed significant change in long-term STBs over time (YRBS; *F =* 4.40, *p* = .011). Scores of long-term STBs are displayed in Figure 4, with slightly worsened scores at timepoint 3 (M= 13.3) for the *high-severity* group.

**Figure 4:**
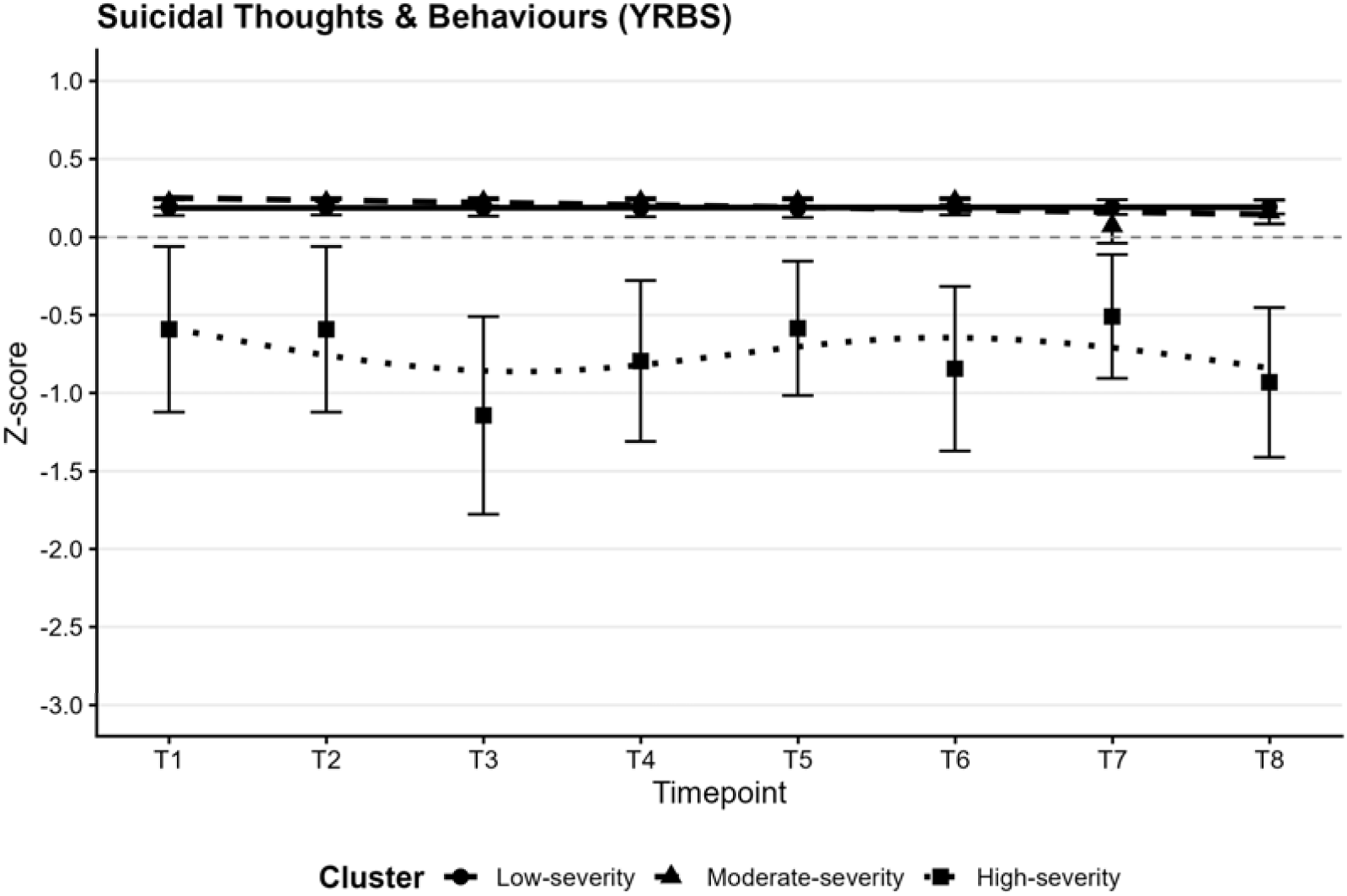
Cluster long-term suicidal thoughts and behaviours (< 12 months) mean standardised scores across early adolescence (T1-8). *Note.* Cluster mean standardised scores (with standard error bars [±1 SE]) and GAMM-smoothed trajectories across timepoints 1-8 for all three clusters for long-term STBs (YRBS). Z-scores were inverted for YRBS to ensure directional alignment across all variables (before inversion, higher scores on the YRBS represented ‘worse’ scores) prior to GAMMs GAMM smooths were estimated using cluster-specific cubic regression splines (k=4) fitted via REML.

## DISCUSSION

This study identified three clusters of suicidality severity (*low-, moderate-* and *high-severity*), which were characterised by differences in psychological distress, lifestyle factors and the prevalence of STBs among adolescents. Psychological distress, sleep, mindfulness, social connectedness, eating habits, and suicidality accounted for 93.1% of the variance between clusters, indicating that lifestyle factors meaningfully distinguish adolescent severity profiles. Relative contributions to group discrimination showed that recent suicidal ideation and sleep quality were the strongest predictors of cluster membership. While other lifestyle factors showed moderate to minimal discriminant contributions, sleep quality emerged as a particularly important correlate differentiating clusters. This is consistent with previous LABS research, finding poor sleep quality to be longitudinally associated with increased suicidal ideation (Crethar et al., 2025).

The *low-severity* cluster displayed low suicidality and psychological distress, highest levels of protective lifestyle indicators (sleep, social connectedness, mindfulness traits, eating habits, and physical activity), the highest wellbeing scores and an absence of long-term STBs. This supports the protective role of positive lifestyle factors, consistent with research indicating that engagement in health-promoting behaviours is associated with reduced suicidality risk (Lee et al., 2024).

The *moderate-severity* cluster reported scores between the other clusters on most outcomes, with the notable exception of suicidality, where scores were comparable to the *low-severity* cluster. This graded pattern is consistent with previous research indicating a relationship between lifestyle factors and mental health outcomes (Grasdalsmoen et al., 2020; Pfledderer et al., 2019). GAMM trajectory analyses indicated that this cluster showed significant change over time for recent suicidal ideation, mindfulness, and eating habits. Declines over time for mindfulness and eating habits, suggest that deterioration in health behaviours may serve as an early indicator of emerging vulnerability in adolescents who do not yet present with elevated suicidality.

The *high-severity* cluster exhibited elevated suicidality and psychological distress; as well as poor sleep, social connectedness, physical activity, mindfulness traits, and eating habits, and reported the lowest SES across groups. Furthermore, this cluster reported the lowest wellbeing scores and highest suicidality across timepoints, with the other two clusters reporting markedly lower scores by comparison. This pattern of consistently poor psychological and lifestyle outcomes presents longitudinal empirical evidence to support previous findings linking psychological distress (Shawon et al., 2023) and lower socioeconomic status (Madigan & Daly, 2023) to adolescent suicidality vulnerability and extends cross-sectional evidence suggesting that deficits across lifestyle domains may compound that risk (Xiao et al., 2021).

A major strength of this study is its longitudinal design. Unlike many studies that rely on cross-sectional methodology or even the quasi-longitudinal design of two waves, this study utilised data collected every four months over a five-year period. The longitudinal design allowed for the identification of clustered, distinct groups based on timepoints 9–15 that had remained relatively stable between the earlier timepoints 1–8. This finding suggests that severity profiles become more differentiated and stable as adolescents progress through developmental stages – a conclusion supported by research indicating that suicide risk typically increases from around age 15 onwards (AIHW, 2025; Glenn et al., 2020). The *high-severity* cluster demonstrated greater variability in suicidality scores compared to the other clusters, revealing less predictability in developmental trajectories. This increased volatility appears to be a distinct feature and could serve as an early warning sign for parents, teachers, and mental health professionals. The fluctuating nature of suicidality in the *high-severity* group aligns with previous research (Platt et al., 2013) that has emphasised the emotion-related impulsivity and suicidality fluctuations throughout adolescence. Of particular importance, is that for the *high-severity* cluster, psychological wellbeing and engagement in health promoting lifestyle factors in early adolescence appeared to weaken with age.

GAMMs confirmed the *high-* and *moderate-severity* clusters had significantly poorer outcomes than the *low-severity* cluster across most variables, while the low-severity cluster remained stable across all timepoints. Notably, the *moderate-severity* cluster did not differ significantly from the *low-severity* cluster on measures of suicidality, suggesting comparable suicidality profiles despite broader differences in wellbeing and lifestyle functioning. The *high-severity* cluster showed the most widespread deterioration, with significant declines in wellbeing, social connectedness, physical activity, eating habits, and STBs. This suggests that vulnerable young people tend to lose protective factors while concurrently experiencing lowered wellbeing and greater suicidality severity. Supporting the critical nature of implementing early interventions at age 12–14 years when initial warning signs appear. The examination of long-term STBs across early adolescence revealed that young people with elevated symptom severity may report suicidality prior to 12 years of age, emphasising the need for earlier screening and preventative interventions in pre-adolescent populations. Suicidality levels remained relatively stable in their rate of change across early adolescence. This suggests that lifestyle and wellbeing factors may be more sensitive to change during this developmental period and represent possible useful targets for early monitoring and intervention.

Several limitations are noted. First, while self-report measures offer important insights into adolescents’ subjective experiences, they are also vulnerable to common method biases such as social desirability (Ridgers et al., 2012). Future studies should incorporate multiple informants (Xiao et al., 2021) or objective measures of lifestyle factors to enhance measurement validity. Second, while the cluster analysis identified meaningful subgroups, there were uneven sample sizes across groups, with the h*igh-severity* group having the lowest sample size. The number of outliers that were windsorised for the *high-*severity cluster exceeded 10% of cases for five variables, limiting generalisability. Larger and more diverse samples could enhance the generalisability of these findings and reveal additional subgroups with distinct severity profiles. Third, analyses were focused on between-group differences over time rather than within-person changes. Consequently, we cannot infer causality between lifestyle factor changes and suicidality fluctuations within the same adolescent over time. Fourth, this study did not examine potential moderating factors such as family dysfunction, which may influence the relationship between lifestyle factors and suicidality. Future research could explore how contextual factors shape severity profiles and developmental trajectories at the individual level over time, as intraindividual variability may represent a clinically meaningful marker of vulnerability.

## CONCLUSION

This study advances our understanding of adolescent suicidality through a longitudinal identification of distinct severity profiles. By using panel data to examine the developmental trajectories of suicidality, psychological distress, and lifestyle factors, a distinct clustering of protective and risk factors emerged over time. Adolescence in the *high-severity* cluster showed elevated suicidality from age 12–17, alongside poorer psychological and lifestyle factors. This indicates that targeted interventions in early adolescence should be considered more broadly. In addition, patterns of sleep, social connection, physical activity, mindfulness, and eating habits clustered together; with different groups showing distinct levels of low, moderate, and high activation of these health promoting behaviours. Greater knowledge of how risk and protective factors cluster together and evolve throughout teenage development can inform more targeted strategies for suicide prevention. Above all, the importance of a proactive and preventative approach to reducing suicidality centred on early intervention cannot be overemphasised.

## Declaration of interests

The authors report no conflicts of interest.

## Data Availability

The datasets generated and analysed during the current study are not publicly available due the fact that they constitute an excerpt of research in progress but are available from the corresponding author on reasonable request.

## Ethical Information

Ethical approval for the project was provided on 9 May 2018 by the University of the Sunshine Coast Human Research Ethics Committee (reference A181064), and cohort participants/caregivers provided written informed assent/consent as appropriate.

## Acknowledgements

This project used data from the ongoing Longitudinal Adolescent Brain Study (LABS), run by the Youth Mental Health research team at UniSC’s Thompson Institute. The authors thank the young people who participated in this research and the researchers who have collected data. The authors report financial support by the Australian Commonwealth Government’s ‘Research Training Program Scholarship’ and the ‘Prioritising Mental Health Initiative (2018–25)’. In addition, the authors report financial support by a Queensland Mental Health Commission’ grant under Every life: The Queensland Suicide Prevention Plan 2019-2029. The authors declare that they have no competing or potential conflicts of interest.

## Notes

### Competing Interest Statement

The authors have declared no competing interest.

### Author Declarations

The University of the Sunshine Coast Human Research Ethics Committee gave ethical approval for this work.

