## Supplementary material for "Developmental trajectories of suicidality and lifestyle factors across adolescent severity profiles": Figures S1-S2

Figure S2. Cluster analysis scree plot

|  | Coefficients |
| --- | --- |
| 1 | 2577.14 |
| 2 | 1593.67 |
| 3 | 1313.11 |
| 4 | 1156.87 |
| 5 | 1048.42 |
| 6 | 944.938 |
| 7 | 866.529 |
| 8 | 788.391 |
| 9 | 723.01 |
| 10 | 662.193 |
| 11 | 611.479 |
| 12 | 564.6 |
| 13 | 521.416 |
| 14 | 483.057 |
| 15 | 449.85 |
| 16 | 422.407 |
| 17 | 395.61 |
| 18 | 373.953 |
| 19 | 352.87 |
| 20 | 333.421 |
