## Supplementary figures and images for "Developmental trajectories of suicidality and lifestyle factors across adolescent severity profiles"

### Figures S1-S2

Figure S1. Cluster analysis dendrogram

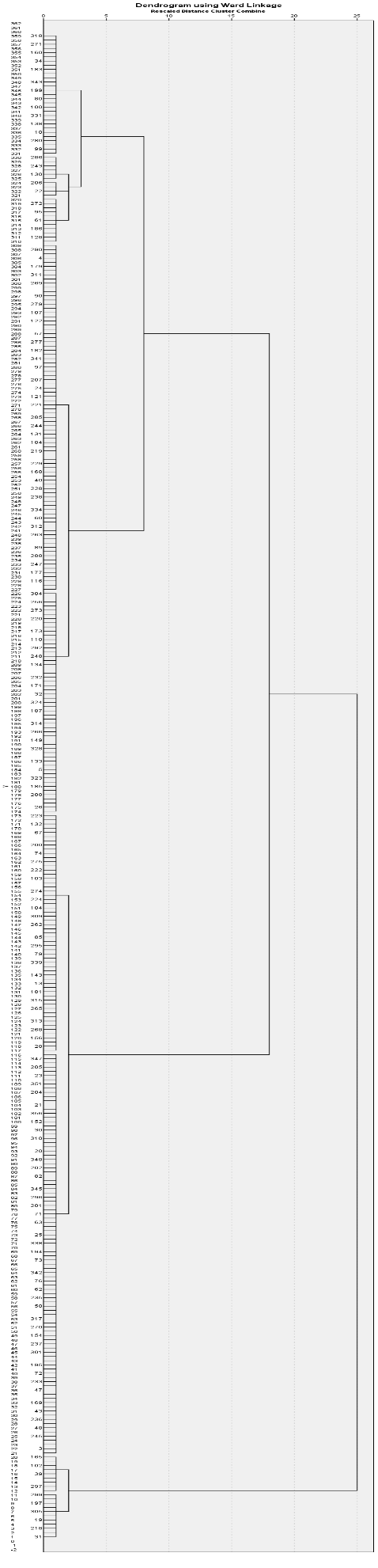
