## Supplementary material for "Developmental trajectories of suicidality and lifestyle factors across adolescent severity profiles": Table S1

Table S1. Mean Z-scores and standard deviation for suicidality, psychological distress, and lifestyle factor variables across the three clusters with corresponding between-clusters ANOVA results.

| Outcome |  | | | ANOVA F *(p)* | Effect sizes, *d* | | |
| --- | --- | --- | --- | --- | --- | --- | --- |
|  | *Low-severity*   (*n*  = 43) | *Moderate-severity*   (*n* = 37) | *High-severity*   (*n* = 18) | *F* [*df*=2, 358] | Low- vs. High-severity | Low- vs. Moderate-severity | High- vs. Moderate-severity |
| Suicidal Ideation (SIDAS) | 0.29 (0.15) | 0.28 (0.16) | -0.89 (1.31) | 110.5 (<.001) | 1.65† | 0.06 | -1.55† |
| Psychological Distress (K10) | 0.71 (0.38) | -0.04 (0.63) | -1.36 (0.83) | 298.5 (<.001) | 3.77† | 1.47† | -1.88† |
| Sleep (PSQI) | 0.70 (0.52) | -0.06 (0.67) | -1.37 (0.83) | 247.9 (<.001) | 3.31† | 1.28† | -1.81† |
| Social Connectedness (SCS) | 0.67 (0.49) | -0.16 (0.76) | -1.10 (1.10) | 142.1 (<.001) | 2.46† | 1.32† | -1.06† |
| Mindfulness (MAAS-A) | 0.87 (0.47) | -0.42 (0.61) | -1.03 (0.93) | 264.8 (<.001) | 2.98† | 2.39† | -0.84† |
| Eating Habits (FFQ) | 0.72 (0.56) | -0.40 (0.67) | -0.74 (1.20) | 118.5 (<.001) | 1.83† | 1.83† | -0.39‡ |
| Physical Activity (MVPA) | 0.52 (0.79) | -0.18 (0.92) | -0.75 (0.96) | 56.2 (<.001) | 1.51† | 0.82† | -0.61‡ |

*Note.* Cohen's *d* effect sizes (*d*): †Large effect (*d* > 0.8); ‡Medium effect (*d* > 0.5). ANOVA = analysis of variance.
