## Supplementary material for "Developmental trajectories of suicidality and lifestyle factors across adolescent severity profiles": Table S2

Table S2. Prevalence of YRBS suicide-related behaviours.

| YRBS Item | *Low-severity*  (n = 43) | *Moderate-severity*  (*n* = 37) | *High-severity*  (*n* = 18) |
| --- | --- | --- | --- |
| Considered attempting suicide | | | |
| Yes | 0 (0%) | 1 (2.70%) | 9 (50.00%) |
| No | 43 (100%) | 35 (94.59%) | 9 (50.00%) |
| Missing | - | 1 (2.70%) | - |
| Made a suicide plan | | | |
| Yes | 0 (0%) | 1 (2.70%) | 6 (33.33%) |
| No | 43 (100%) | 35 (94.59%) | 12 (66.67%) |
| Missing | - | 1 (2.70%) | - |
| Attempted suicide | | | |
| No attempt | 0 (0%) | 36 (97.30%) | 14 (77.78%) |
| ‘one or more attempts’ | 43 (100%) | 0 | 4 (22.22%) |
| Missing | - | 1 (2.70%) | - |

*Note.* The YRBS assesses STBs over the past 12 months. Where participants attended multiple timepoints, they were allocated to ‘considered attempting suicide’, ‘made a suicide plan’ or ‘attempted suicide’ if at least once over T9-15 they responded ‘yes’ or reported one or more suicide attempt.
