## Supplementary material for "Developmental trajectories of suicidality and lifestyle factors across adolescent severity profiles": Table S3

Table S3. Generalised Additive Mixed Model (GAMM) results for nine outcomes across timepoints 1-8 by cluster.

| Outcome | Parametric Effects (vs. Low-severity) | | | | Smooth Terms s(tp) by Cluster | | | Model Fit | |
| --- | --- | --- | --- | --- | --- | --- | --- | --- | --- |
|  | *High-severity* | | *Moderate-severity* | | *Low-severity* | *Moderate-severity* | *High-severity* | *R²*adj | Dev. Exp. (%) |
|  | *b* | *p* | *b* | *p* | edf / *F / p* | edf / *F / p* | edf / *F / p* |  |  |
| Sleep (PSQI) | −1.56 | < .001 | −0.74 | < .001 | 1.39 / 2.44 / .062 | 2.49 / 1.20 / .211 | 1.00 / 0.70 / .404 | 0.68 | 73.0 |
| Suicidal Ideation (SIDAS) | −0.76 | < .001 | −0.21 | .098 | 1.00 / 0.50 / .481 | 2.57 / 5.65 / .006 | 1.00 / 0.17 / .679 | 0.40 | 48.8 |
| Psychological Distress (K10) | −1.47 | < .001 | −0.80 | < .001 | 1.00 / 0.34 / .562 | 1.00 / 1.57 / .210 | 2.16 / 1.86 / .285 | 0.61 | 67.2 |
| Wellbeing (COMPAS-W) | −1.41 | < .001 | −0.59 | .002 | 1.00 / 1.04 / .309 | 1.00 / 1.81 / .179 | 1.00 / 8.51 / .004 | 0.73 | 77.6 |
| Social Connectedness (SCS) | −1.30 | < .001 | −0.75 | < .001 | 1.00 / 0.71 / .400 | 1.00 / 2.30 / .130 | 1.00 / 4.76 / .030 | 0.66 | 71.6 |
| Physical Activity (MVPA) | −1.12 | < .001 | −0.58 | .003 | 1.00 / 0.00 / .948 | 1.00 / 0.41 / .525 | 1.00 / 17.00 / < .001 | 0.73 | 77.3 |
| Mindfulness (MAAS-A) | −1.38 | < .001 | −0.79 | < .001 | 2.02 / 2.29 / .110 | 1.00 / 18.86 / < .001 | 1.00 / 3.32 / .069 | 0.69 | 73.7 |
| Eating Habits (FFQ) | −1.01 | < .001 | −0.82 | < .001 | 1.41 / 1.87 / .243 | 1.00 / 22.82 / < .001 | 1.31 / 4.25 / .016 | 0.78 | 82.0 |
| Suicidal Thoughts & Behaviours (YRBS) | −0.83 | < .001 | −0.07 | .563 | 1.00 / 0.77 / .382 | 1.00 / 0.01 / .926 | 1.77 / 4.40 / .011 | 0.62 | 67.8 |

*Note.* Models included cluster as a parametric factor (low-severity as the reference group), cluster-specific smooth terms s(tp) for timepoint trajectory (*k*=4, cubic regression spline), and a random effect for participant (s(subj_id), bs = “re”) to account for repeated measures. Parametric coefficients (*b*) represent overall differences between clusters relative to the low-severity cluster. Smooth terms are presented as estimated degrees of freedom (edf), *F-*statistics and *p*-values. Edf values of > 1 indicate non-linear trajectories. All models included a significant random effect of participant (*p <* .001; not shown). All outcomes were standardised (z-scores); inverted outcomes (PSQI, SIDAS, K10, YRBS) were coded such that higher values indicate better functioning. Dev. Exp. = deviance explained.
